# Bayesian network structure for predicting two-year survival in patients diagnosed with non small cell lung cancer

**DOI:** 10.1101/2023.01.27.23284249

**Authors:** Biche Osong, Andre Dekker, Leonard Wee, Johan van Soest, Inigo Bermejo

**Affiliations:** GROW School for Oncology and Developmental Biology, Maastricht University Medical Centre+, Maastricht, the Netherlands

## Abstract

**Purpose:** The aim of this study was to develop and internally validate a clinically plausible Bayesian network structure to predict two-year survival in patients diagnosed with non-small cell lung cancer (NSCLC) and primarily treated with (chemo) radiation therapy by combining expert knowledge and a learning algorithm.

**Summary of background:** The incidence of lung cancer has been increasing. Healthcare providers are trying to acquire more knowledge of the disease’s biology to treat their patients better. However, the information available is more than humans can efficiently process. Predictive models such as Bayesian networks, which can intricately represent causal relations between variables, are suitable structures to model this information. However, commonly known methods for developing Bayesian network structures are limited in healthcare.

**Patients and Methods:** 545 NSCLC patients treated primarily with (chemo) radiation therapy from Maastro clinic in the Netherlands between 2010 to 2013 were considered to develop this Bayesian network structure. All continuous variables were discretized before analysis. Patients with missing survival status and variables with more than 25% missing information were excluded. The causal relationships (arcs) between variables in the data were determined using the hill-climbing algorithm with domain experts’ restrictions. The learning algorithm was run on a number of bootstrapped samples (B=400) and for the final structure, we kept the arcs present in at least 70% of the learned structures. Performance was assessed by computing the area under the curve (AUC) values and producing calibration plots based on a 5-fold cross-validation. In addition, an adapted pre-specified expert structure was compared with a structure developed from the method in this study.

**Results:** Tumor load was included in the main structure due to its high percentage (37%) of missingness and lack of added value. The final cohort used to develop the structure was reduced to 499, excluding 46 (8.4%) patients with missing survival status. The resulting structure’s mean AUC and confidence interval to predict two-year survival was 0.614 (0.499 - 0.730). The AUC of the compared structures was only slightly above the chance level, but the structure based on the method in this study was clinically more plausible.

**Conclusion:** The results of this study show that Bayesian network structures which combine expert knowledge with a rigorous structure learning algorithm produce a clinically plausible structure with optimal performance.

## Introduction

Lung cancer is the second most common cancer and the leading cause of cancer morbidity and mortality in men and second for women after breast cancer worldwide, with non-small cell lung cancer (NSCLC) accounting for approximately 85% of this disease [1, 2]. These statistics actuate healthcare providers to acquire more knowledge and understand the patient condition and disease characteristics for better patient management and treatment outcomes.

However, the amount of information that needs to be processed to ascertain if a patient will experience an event of interest can be challenging even for domain experts [3, 4]. Furthermore, it has been shown that even experienced domain experts specialized in the treatment of lung cancer have limited to no capabilities for predicting patients’ outcomes vis-a-vis prediction models [5–8]. Predictive models such as Bayesian networks (BN), which can structurally represent a domain of interest by causally mapping the domain’s variables, may be more suitable for modeling such information.

Bayesian network structures are either expert(s) specified or algorithm-based [9–11]. However, these methods are limited in a clinical setting due to implausible casual relationships for algorithmic structures or bias for expert structures based on their experience and domain knowledge. Our prior work, which compared the performance of structures from these two sources, showed that algorithm-based structures perform relatively better but with little or no clinical interpretability [12]. On the other hand, expert structures are more clinically interpretable but with relatively inferior performance. Therefore, this study aims to develop a Bayesian network structure that stems from both methods to predict two-year survival in patients diagnosed with lung cancer primarily treated with radiation therapy.

We hypothesize that a symbiotic relationship between domain experts and a robust learning algorithm (expert-algorithm) would yield a clinically interpretable and plausible Bayesian network structure to predict two-year survival for lung cancer patients with optimal performance.

## Materials and methods

We retrospectively collected data of 545 non-small cell lung cancer (NSCLC) patients diagnosed between 2010 to 2013 and eligible for (chemo) radiotherapy treatment at Maastro Clinic, Maastricht, The Netherlands. Patients’ demographics and clinical information such as age, gender, WHO performance status, TNM stage, tumor load, FEV, smoking status, chemotherapy type, and two-year survival status were extracted to establish the Bayesian network.

### Bayesian network

Bayesian networks model the relationships between a set of variables. These relationships are represented in a directed acyclic graph (DAG), where each node in the graph signifies a variable [9].The direction of the link between nodes represents the influence dependency from the causal variable known as the parent node to the affected variable called the child node. Therefore, each variable can be a child or parent to numerous variables, but the process should not contain any loops. In other words, tracing the parent-to-child link should not connect a variable with itself or a variable functioning as a child and parent to another variable [11]. The conditional probability table (CPT) represents the probabilities of each possible state of a node, given the states, its parent node may take [9, 10, 13].

### Structure learning

The structure learning process was bootstrapped (B=400) with varying sample sizes at each run using the hill-climbing algorithm to identify the causal relationships (arcs) between variables in the dataset. Arc strength was evaluated as the rate of occurrence over all the bootstrap runs, and only arcs with an occurrence rate above 70% were included in the final structure. Domain knowledge from multiple experts in the field was employed to restrict the algorithm from forming arcs in clinically implausible directions (so-called blacklist) like age having a causal influence on gender (Table S1 in the supplementary materials). The pseudocode for the expert-algorithm method is outlined in algorithm 1.

#### Algorithm 1

**Expert-Algorithm pseudocode**

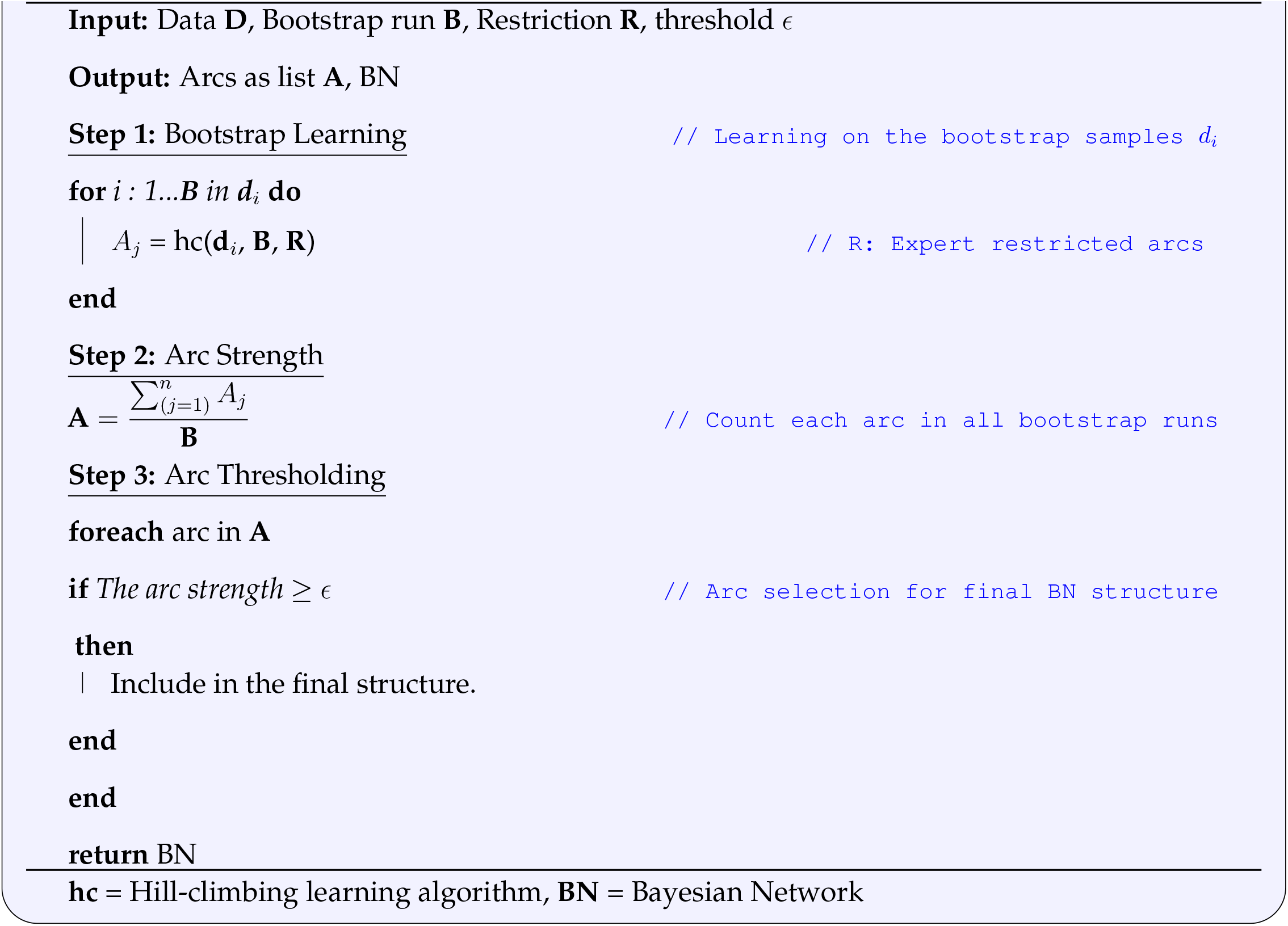

### Statistical analysis

All analysis was conducted in R version 4.1.0 [14] using the bnlearn package [15] and GeNIe a Graphical Network Interface application [16] was used to visualize the developed Bayesian network structures. Tumorload and age were categorized into three groups with cutoff values at the 25th and 75th percentile and the force expiratory volume (FEV) was categorized based on experts opinions. Missing values were imputed using the Multivariate Imputation via Chained Equations (MICE) package [17]. Patients with missing survival status and variables with more than 25% missingness were excluded from the analysis. The predictive performance of the resulting Bayesian network structure was assessed by computing the area under the curve (AUC) using a 5-fold cross validation technique and generating calibration plots.

The main structure was updated with the excluded variable using the **structural.em** function in the bnlearn package to check if the excluded variables having above 25% missingness were crucial. The function learns a Bayesian network from a dataset containing missing information by first inputting the missing data using the expectation-maximization (EM) algorithm and then finds the best possible structure based on the imputed data. The arcs of the main structure were used as a whitelist in the structure update process.

## Results

From the 545 patients in this study, 46 (8.4%) with a missing two-year survival status were excluded from this analysis, reducing the cohort to 499. Tumorload was excluded from further analysis due to its high percentage (37%) of missing information. The median age of patients in this study was 68 (33 - 89). Most of the patients in this study are ex-smokers, with the number of males almost twice that of females. Table 1 shows a detailed descriptions of patients’ characteristics for this study cohort.

**Table 1.**
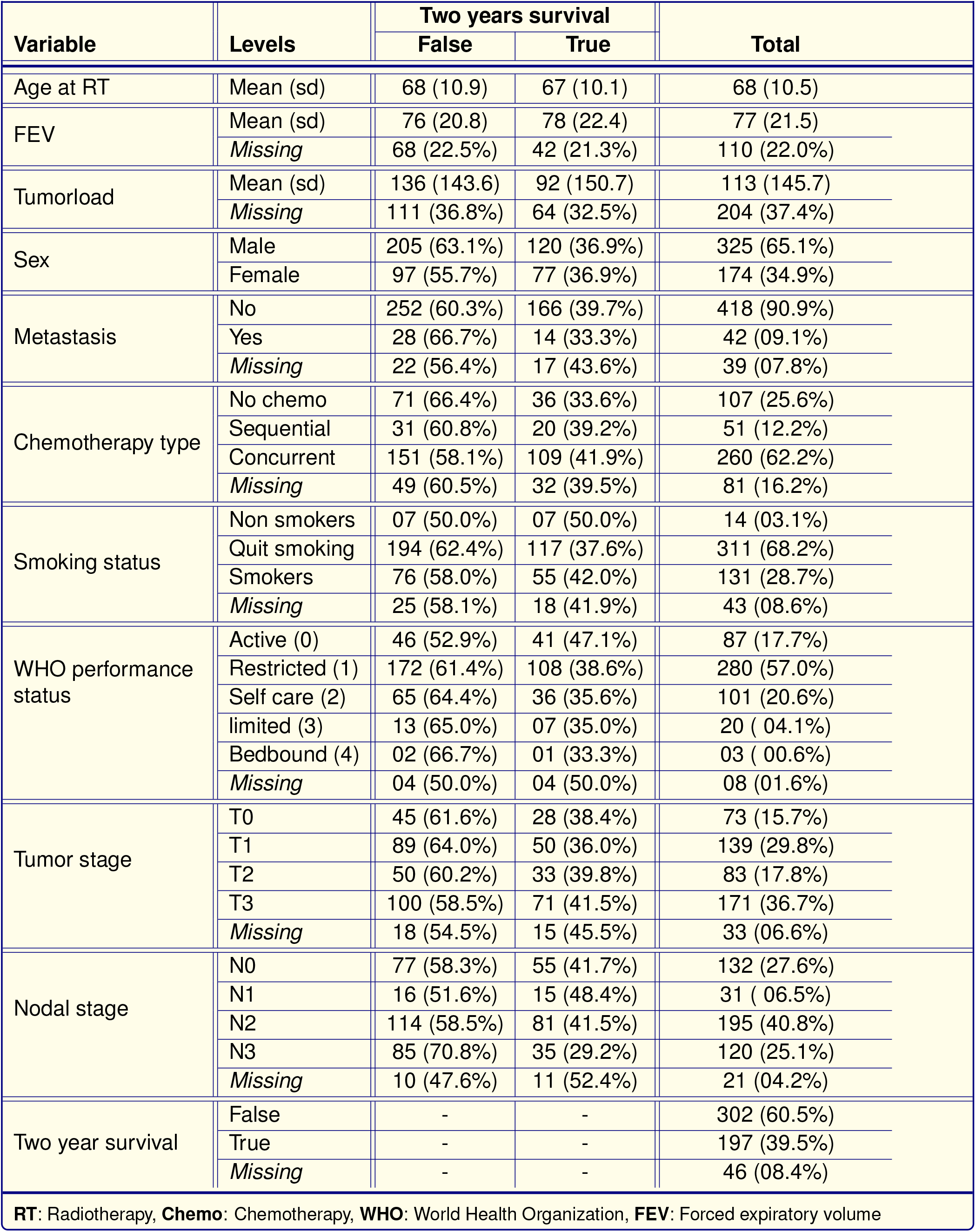
Overview of patient demographics and clinical characteristics

|  |  |  |  |  |
| --- | --- | --- | --- | --- |
| Variable | Levels | Two years survival |  | Total |
|  |  | False | True |  |
| Age at RT | Mean (sd) | 68 (10.9) | 67 (10.1) | 68 (10.5) |
| FEV | Mean (sd) | 76 (20.8) | 78 (22.4) | 77 (21.5) |
|  | Missing | 68 (22.5%) | 42 (21.3%) | 110 (22.0%) |
| Tumorload | Mean (sd) | 136 (143.6) | 92 (150.7) | 113 (145.7) |
|  | Missing | 111 (36.8%) | 64 (32.5%) | 204 (37.4%) |
| Sex | Male | 205 (63.1%) | 120 (36.9%) | 325 (65.1%) |
|  | Female | 97 (55.7%) | 77 (36.9%) | 174 (34.9%) |
| Metastasis | No | 252 (60.3%) | 166 (39.7%) | 418 (90.9%) |
|  | Yes | 28 (66.7%) | 14 (33.3%) | 42 (09.1%) |
|  | Missing | 22 (56.4%) | 17 (43.6%) | 39 (07.8%) |
| Chemotherapy type | No chemo | 71 (66.4%) | 36 (33.6%) | 107 (25.6%) |
|  | Sequential | 31 (60.8%) | 20 (39.2%) | 51 (12.2%) |
|  | Concurrent | 151 (58.1%) | 109 (41.9%) | 260 (62.2%) |
|  | Missing | 49 (60.5%) | 32 (39.5%) | 81 (16.2%) |
| Smoking status | Non smokers | 07 (50.0%) | 07 (50.0%) | 14 (03.1%) |
|  | Quit smoking | 194 (62.4%) | 117 (37.6%) | 311 (68.2%) |
|  | Smokers | 76 (58.0%) | 55 (42.0%) | 131 (28.7%) |
|  | Missing | 25 (58.1%) | 18 (41.9%) | 43 (08.6%) |
| WHO performance status | Active (0) | 46 (52.9%) | 41 (47.1%) | 87 (17.7%) |
|  | Restricted (1) | 172 (61.4%) | 108 (38.6%) | 280 (57.0%) |
|  | Self care (2) | 65 (64.4%) | 36 (35.6%) | 101 (20.6%) |
|  | limited (3) | 13 (65.0%) | 07 (35.0%) | 20 ( 04.1%) |
|  | Bedbound (4) | 02 (66.7%) | 01 (33.3%) | 03 ( 00.6%) |
|  | Missing | 04 (50.0%) | 04 (50.0%) | 08 (01.6%) |
| Tumor stage | T0 | 45 (61.6%) | 28 (38.4%) | 73 (15.7%) |
|  | T1 | 89 (64.0%) | 50 (36.0%) | 139 (29.8%) |
|  | T2 | 50 (60.2%) | 33 (39.8%) | 83 (17.8%) |
|  | T3 | 100 (58.5%) | 71 (41.5%) | 171 (36.7%) |
|  | Missing | 18 (54.5%) | 15 (45.5%) | 33 (06.6%) |
| Nodal stage | N0 | 77 (58.3%) | 55 (41.7%) | 132 (27.6%) |
|  | N1 | 16 (51.6%) | 15 (48.4%) | 31 ( 06.5%) |
|  | N2 | 114 (58.5%) | 81 (41.5%) | 195 (40.8%) |
|  | N3 | 85 (70.8%) | 35 (29.2%) | 120 (25.1%) |
|  | Missing | 10 (47.6%) | 11 (52.4%) | 21 (04.2%) |
| Two year survival | False | - | - | 302 (60.5%) |
|  | True | - | - | 197 (39.5%) |
|  | Missing | - | - | 46 (08.4%) |
| RT: Radiotherapy, Chemo: Chemotherapy, WHO: World Health Organization, FEV: Forced expiratory volume |  |  |  |  |

The variable age was discretized into three groups with cutoff values at the 25th and 75th percentile (Figure S2 supplementary materials). Patients between the cutoff values are considered elderly as shown in equation 1 while patients below and above the cut-off values were considered adults and seniors, respectively. The forced expiratory volume (fev) was also discretized into four groups based on experts suggestion as shown in equation 2.

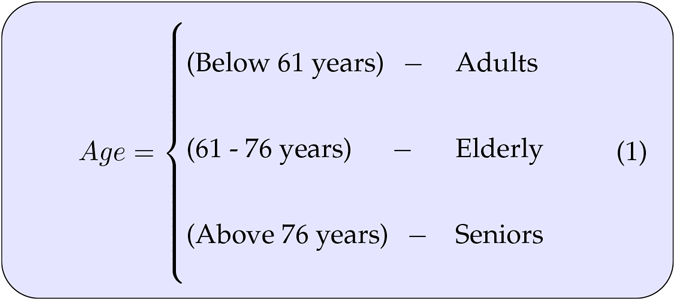

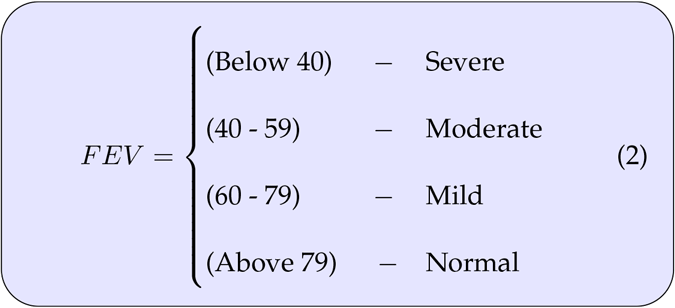

The WHO performance status was recategorized into four groups by combining patients in the limited (3) and bed-bound (4) categories into the same category (bed-bound) because of the very low number of patients in the two categories (Table 1). Also, they both have similar characteristics (See Table S2 in the supplemental material for further explanations).

Diagnostic plots were created to ensure that the imputations has converged to the desired distribution. The convergence check plot (Figure S1 supplemental material) of the imputed values suggest the imputation has converged to the target distribution. Furthermore, the density plot (Figure S4 supplemental material) which compare the distribution of the imputed and observed values confirms that the imputations are reasonable since the distribution of the imputed and observed values are very similar.

Table 2 shows the results of the aggregated arcs from the bootstrap structure learning process using the hillclimbing (hc) algorithm and expert restriction (Blacklist). The arc strength shows the percentage of occurrence of each arc in the bootstrap runs.

**Table 2.** Bootstrap run output with arcs and strength for the main structure.

| Arcs Dependencies (A) |  | Strength |
| --- | --- | --- |
| Parent node | Child node |  |
| Gender | Two year survival | 0.65 |
| Gender | FEV | 0.64 |
| **WHO PS** | **Two year survival** | **0.70** |
| WHO PS | Chemotherapy type | 0.43 |
| Age | Two year survival | 0.61 |
| **Age** | **WHO PS** | **0.84** |
| **Age** | **Chemotherapy type** | **0.99** |
| **Tumor stage** | **Two year survival** | **0.73** |
| Tumor stage | WHO PS | 0.43 |
| **Tumor stage** | **Nodal stage** | **0.91** |
| Tumor stage | FEV | 0.64 |
| **Tumor stage** | **Chemotherapy type** | **0.94** |
| **Nodal stage** | **Two year survival** | **0.84** |
| **Nodal stage** | **WHO PS** | **0.91** |
| **Nodal stage** | **FEV** | **1.00** |
| **Nodal stage** | **Chemotherapy type** | **1.00** |
| Metastasis | Two year survival | 0.42 |
| **Metastasis** | **WHO PS** | **0.92** |
| Metastasis | FEV | 0.66 |
| **Metastasis** | **Chemotherapy type** | **0.76** |
| FEV | Two year survival | 0.63 |
| **FEV** | **WHO PS** | **0.86** |
| **FEV** | **Chemotherapy type** | **0.97** |
| Chemotherapy type | Two year survival | 0.34 |
| **Smoking status** | **Two year survival** | **0.86** |
| **Smoking status** | **FEV** | **0.82** |

FEV: Forced expiratory volume, WHO PS: World Health Organization performance status

A threshold of 0.7 was chosen to decide which arcs should be included or excluded in the Bayesian network structure. A higher threshold value ensures that the conditional probability table (CPT) of the outcome does not grow too large, which can cause the structure to overfit. Therefore, the chosen threshold helps restrain the structure from overfitting but allows enough room for structural complexity for optimal performance.

The resulting Bayesian network structure based on the chosen threshold is presented in figure 1. Of the 26 arcs produced during the bootstrap structure learning process, only 15 had an arc strength above the selected threshold and were used to develop the Bayesian network structure. Four variables directly influenced the response of interest (gray arcs), while gender was isolated on the structure because it was neither a parent nor a child to any variable in the network.

**Fig 1.**
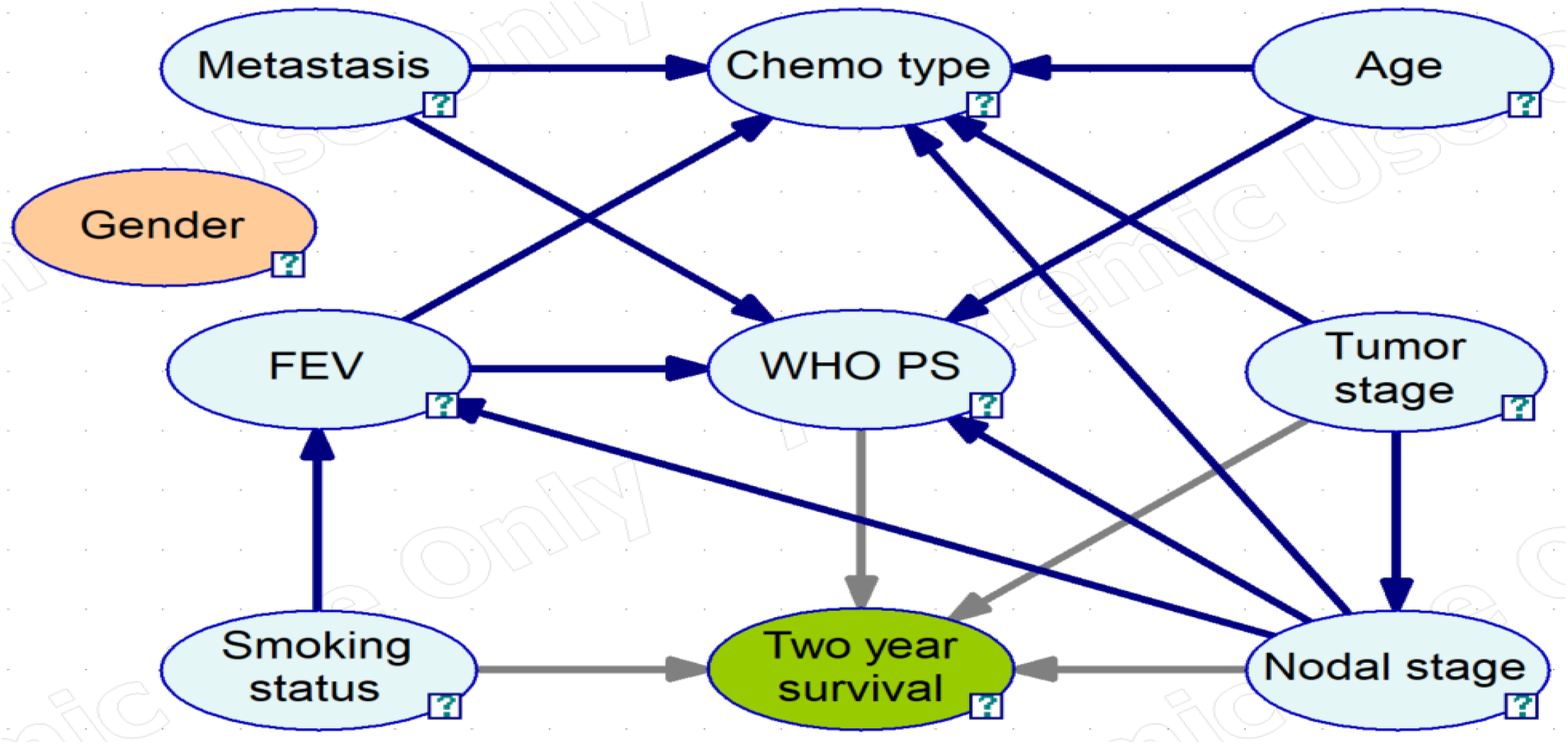
Resulting Bayesian network structure to predict two-year survival from the expert-algorithm method. The oval structure represent the variables (Node), and the arrows indicate the direction of the causal-effect relationships. Grey arrows indicate a direct parental link to the outcome of interest **FEV** = Forced expiratory volume, **Chemo** = Chemotherapy, **WHO PS** = World Health Organization performance status

Figure 2 shows the performance assessment results when the resulting Bayesian network structure was used to predict two years survival in lung cancer patients using the repeated (r=50) 5-fold cross validation technique. The left figure shows the area under the curve and confidence intervals of the respective folds with a mean value of 0.614 (0.499 - 0.730). The right figure gives a measure of how similar the predicted probabilities are to the observed probabilities with calibration assessed in term of the degree of deviation of the points (color) from the 45 degree line (dotted gray).

**Fig 2.**
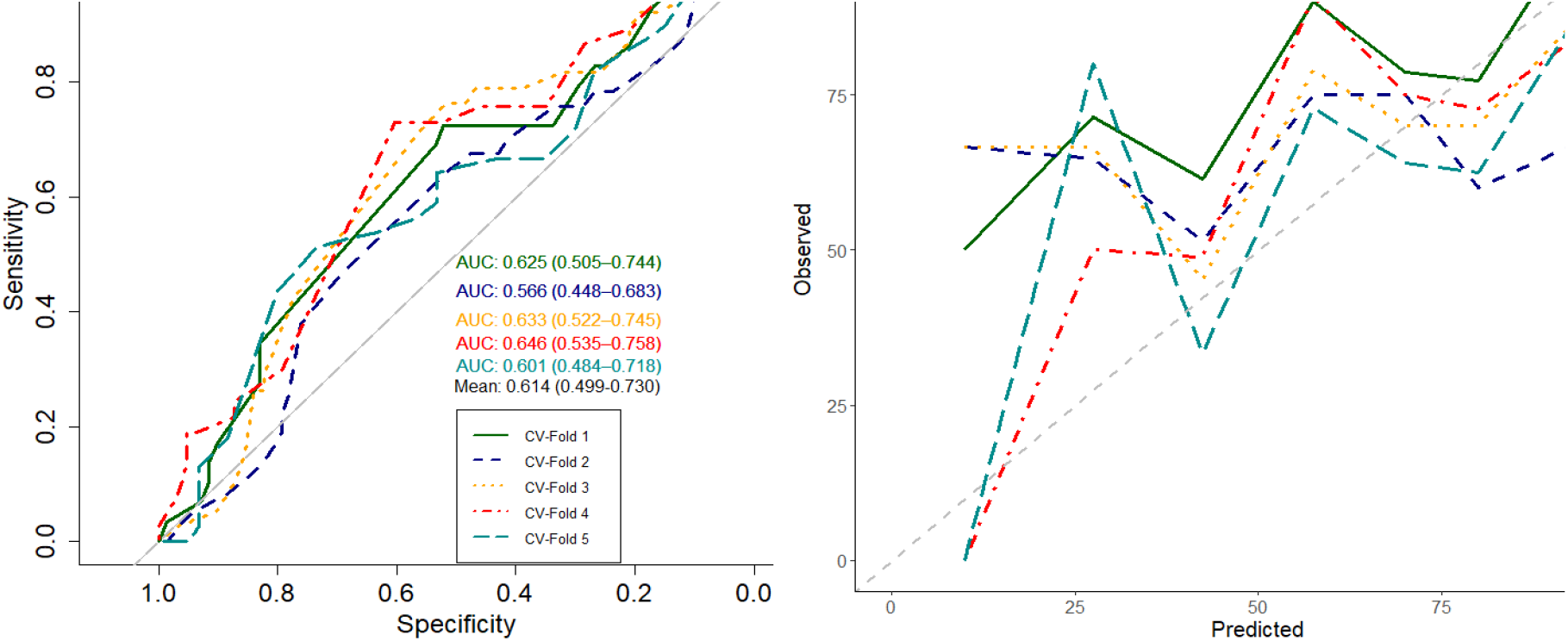
The area under the curve and calibration plot of the structure for predicting two-years survival.

Updating the main structure with the excluded variable included three arcs (red) between **tumor load** and **tumor stage, tumor load** and the presence of **metastasis**, the presence of **metastasis** and **two years survival** (Figure S3 in the supplemental material). However, the addition of these arcs had no significant improvement on the mean AUC value. Gender was again not connected to any other variable in the structure.

To assess the expert-algorithm method, the structure and performance of an expert pre-specified structure from Jochems et al. [18] was compared with a structure resulting from the expert-algorithm procedure with a threshold of 0.7 and the same variables. The aggregated arcs results from the bootstrap runs based on these variables are presented in table 3

**Table 3.** Bootstrap run output with arcs and strength for structure comparison.

| Arcs Dependencies (A) |  | Strength |
| --- | --- | --- |
| Parent node | Child node |  |
| WHO PS | Two year survival | 0.43 |
| Age | Two year survival | 0.38 |
| **Age** | **WHO PS** | 0.94 |
| Tumor stage | Two year survival | 0.36 |
| Tumor stage | WHO PS | 0.10 |
| Tumor stage | Nodal stage | 0.68 |
| **Nodal stage** | **Two year survival** | 0.74 |
| **Nodal stage** | **WHO PS** | 0.92 |
| WHO PS: World Health Organization performance status |  |  |

The structure from Jochems et al. [18] was adapted in this analysis because of the missing total tumor dose variable. Figure 3 shows the adapted structure from Jochems et al. [18] and that resulting from the expert-algorithm method respectively. The only similarity in the structures is that the outcome has just one parent but the variables are completely different with the adapted structure having WHO performance score as parent and nodal stage for the expert-algorithm structure. Based on the differences, the expert-algorithm structure uses one variable and arc less with the outcome being an end node (having no child) compared to the adapted structure.

**Fig 3.**
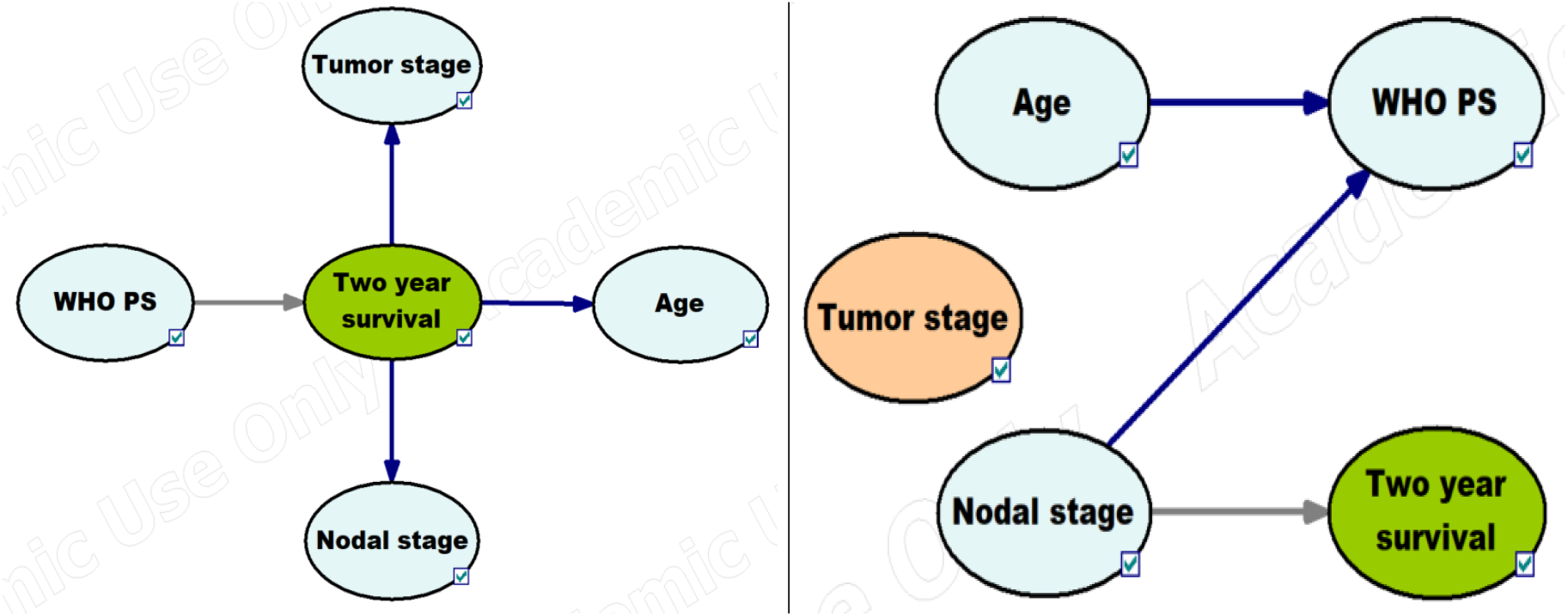
Adapted structure from Jochems et al. [18] and that from the expert-algorithm procedure respectively. The oval structure represents the variables (Node), and the arrows indicate the direction of the causal-effect relationships. Grey arrows indicate a direct parental link to the outcome of interest **WHO PS**: World Health Organization performance status

The performance of both structures was only slightly better than flipping a coin (Figure S5 supplemental material) with an area under the curve of 0.56 (0.517 - 0.613) for the expert-algorithm structure and 0.53 (0.489 - 0.582) for the adapted structure from Jochems et al. [18]. Though both structures had poor performance with just one arc to the outcome, the expert-algorithm structure had a slightly higher discriminating ability than the adapted structure, but this difference was not statistically significant (p-value = 0.413).

To further evaluate the performance of the structures, their respective calibration plots were produced and overlaid (Figure S5 supplemental material). They show that both plots were poorly calibrated given how distant the points are from the diagonal dotted gray line. However, the expert-algorithm structure is better calibrated relatively with more points closer to the diagonal line.

## Discussion

We have developed a novel structure learning method to produce clinically plausible and interpretable Bayesian networks resulting from an interplay of a learning algorithm and experts’ restrictions. The structure produced due to our expert-algorithm method was used to predict two-year survival in NSCLC patients. A resulting structure from the application of our expert-algorithm method was compared to an adapted expert pre-specified structure. Both structures produced comparable results in terms of AUC and calibration. However, the expert-algorithm structure had a slightly better performance with one variable and arc less than the adapted structure and more clinically plausible arcs.

Numerous Bayesian network structures have been developed to predict survival in lung cancer patients [19], and some of these structures stem from our group [12, 18, 20–22]. These structures were either inferred from the data by a structure learning algorithm or pre-specified by expert(s). Jochems et al. [18] has even compared the performance of structures derived from both methods and showed that expert-based structures were more performant than algorithm-based structures, although the difference was not statistically significant. To our knowledge, this is the first time a Bayesian network structure has been developed from routine clinical care data that applies both structure learning methods while considering the clinical sanity and interpretability of the resulting structure—in other words, developing a Bayesian network structure which is suited for clinical implementation, because Bayesian network structures which captures domain knowledge are more interpretable which is essential for clinical decision making.

Bayesian network structure learning process can be time-consuming and computer-intensive, especially for expert base and algorithmic structures like the hill-climbing algorithm, which test pairs of variables to determine whether edges should be included or removed from the structure respectively. Therefore applying our expert-algorithm method, which restricts candidate solutions (arcs) from being evaluated during the search process, could significantly improve structure learning in radiotherapy fields, which involves using high dimensional data, as is the case with radiomics studies. Furthermore, our expert-algorithm method could serve as a means to perform variable selection in the structure learning process of Bayesian networks, something missing in literature since variable selection is mainly performed manually by experts. When a high threshold is applied to the bootstrap object, it leads to the removal of arcs with strength smaller than the specified threshold and possibly variables with arc strength inferior to the threshold as in the case for gender in figure 1. Finally, the expert restriction and the use of a threshold ensure that the resulting structure is clinically correct and includes only relevant relationships in the data with optimal performance.

This study hypothesized that combining experts’ knowledge with a learning algorithm would yield a more clinically correct structure. The results comparing the structures from both methods support this hypothesis. Although both structures performed relatively poorly, we observed a significant improvement in our expert-algorithm structure as most of the arcs in the adapted structure were either reversed or modified to clinically acceptable arcs. However, the arc from WHO PS to the outcome, present in the adapted structure, was absent in the expert-algorithm structure. One possible explanation for this difference could be that WHO PS alone is not a good correlate to survival, but its interplay with other variables increases its influence on survival, as seen in the primary structure (Figure 1) with an arc strength of 0.73 as opposed to 0.43 (Table 2 and 3 respectively). Similar conclusions can be drawn from the study by Jayasurya et al. [20] as the arc (WHO PS to Survival) was also present in their structure with a link weight of -0.169, suggesting that WHO PS has limited ability to predict survival correctly.

The structures presented in this study are not intended for clinical use at the moment but to encourage further research on how to merge these two methods best to develop a Bayesian network structure for a more clinically correct structure but with optimal predictive performance. Therefore, this study is not devoid of limitations mainly because of its retrospective nature, which implies the possibility of data bias. Furthermore, the main structure did not include predictive variables such as the number of positive lymph nodes on the PET scan (PLNS) and tumor load. Although tumor load was available as a variable in this study, it was excluded due to missing information. Faehling, Schwenk, Kopp, Fallscheer, Kramberg, and Eckert [23] has shown that tumor load is an essential factor for overall survival, and patients with lower tumor load have a better outcome than patients with larger tumor load. PLNS, on the other hand, was unavailable in this study which makes the structural comparison somewhat unfair. Also, information is lost with the discretization of continuous variables, which might explain the low predictive power of tumor load in this study coupled with the high missingness. Lastly, this study’s threshold selection is set arbitrarily, which means a large threshold will lead to a sparse network that only partially represents the domain, and a small threshold yields a dense network (complex, squiggly, and harder to read) that might overfit the data. Future researchers should focus on finding an optimal threshold that addresses the shortcomings of the present thresholding.

## Conclusion

We have developed a Bayesian network structure from routine clinical data for predicting two-year survival in lung cancer patients treated with (chemo) radiotherapy. Our expert-algorithm method uses bootstrapping with arc restriction in the structure learning process and assesses the robustness of causal relationships. Therefore, selecting the most robust relationships overall bootstrapping samples produces a structure that captures all relevant relationships within the data with a reduced chance of adding spurious links. In the future, we intend to use this method to evaluate different structures learned from different data sets and perform a privacy-preserving distributed learning approach to structure learning in Bayesian networks.

## Data Availability

All data produced in the present study are available upon reasonable request to the authors

## Supplemental materials

**Fig 1.**
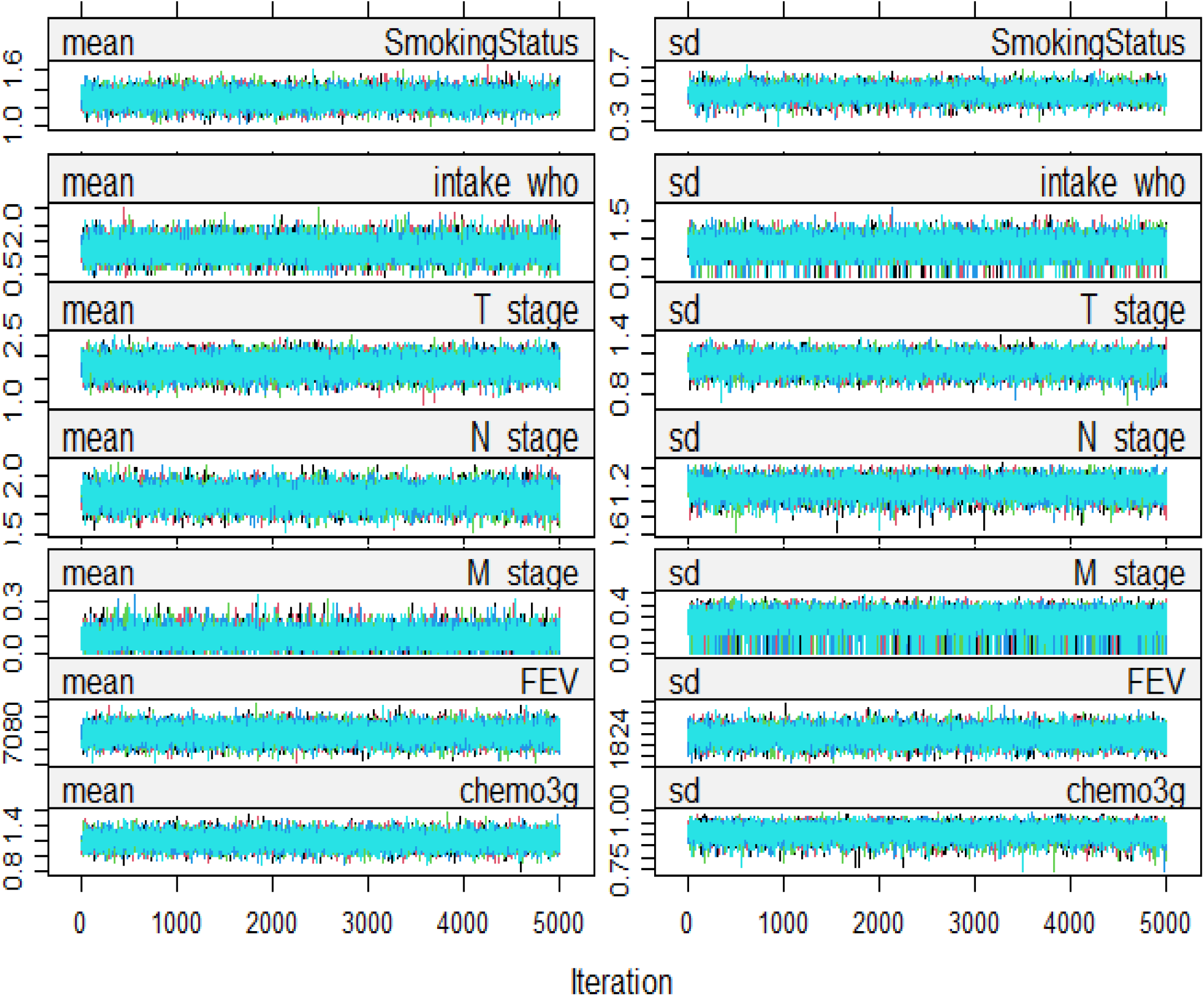
The convergence check plot shows the mean (left) and standard deviation (right) of the imputed values against iteration number. The plot suggests the imputation has converged to the target distribution given the good mix/intermingling of the streams and its trends-free nature at the later part of the iterations

**Fig 2.**
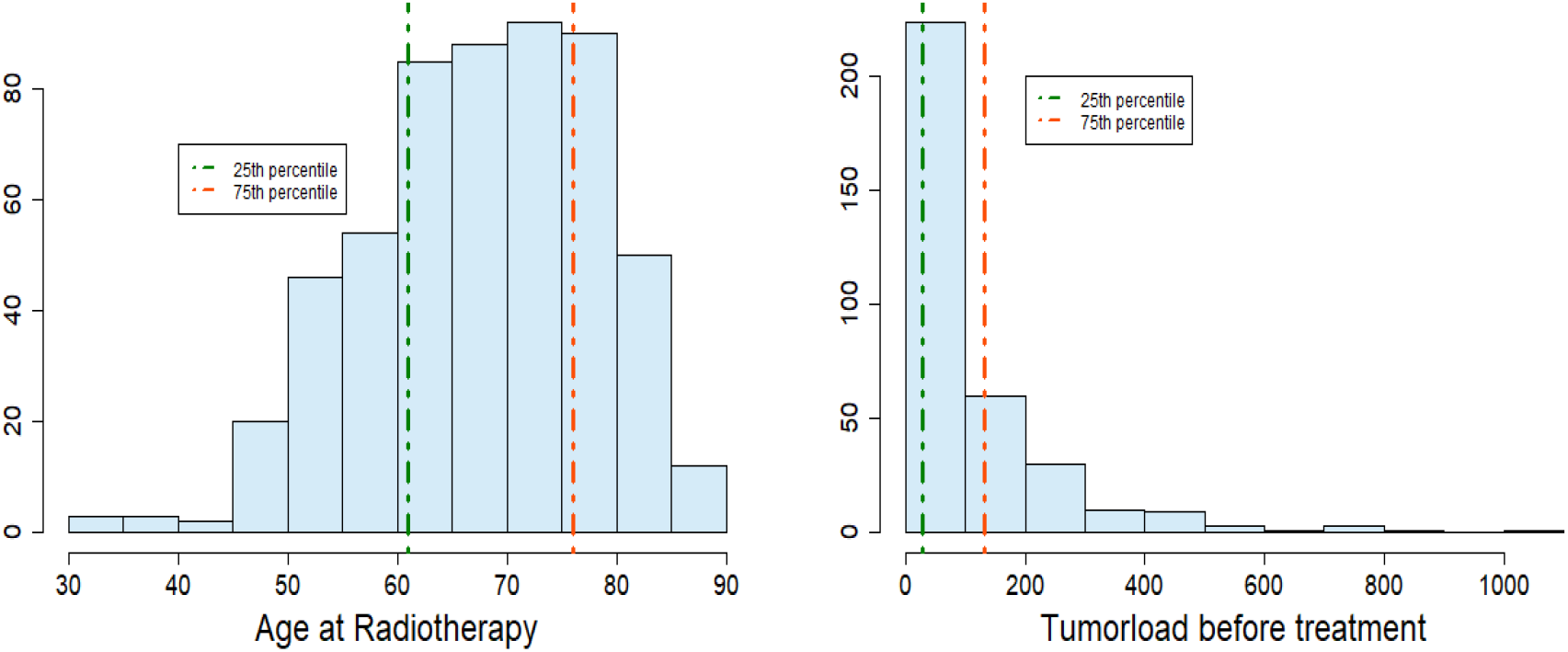
Histogram of age and tumorload with vertical lines at the 25th and 75th percentile respectively.

**Fig 3.**
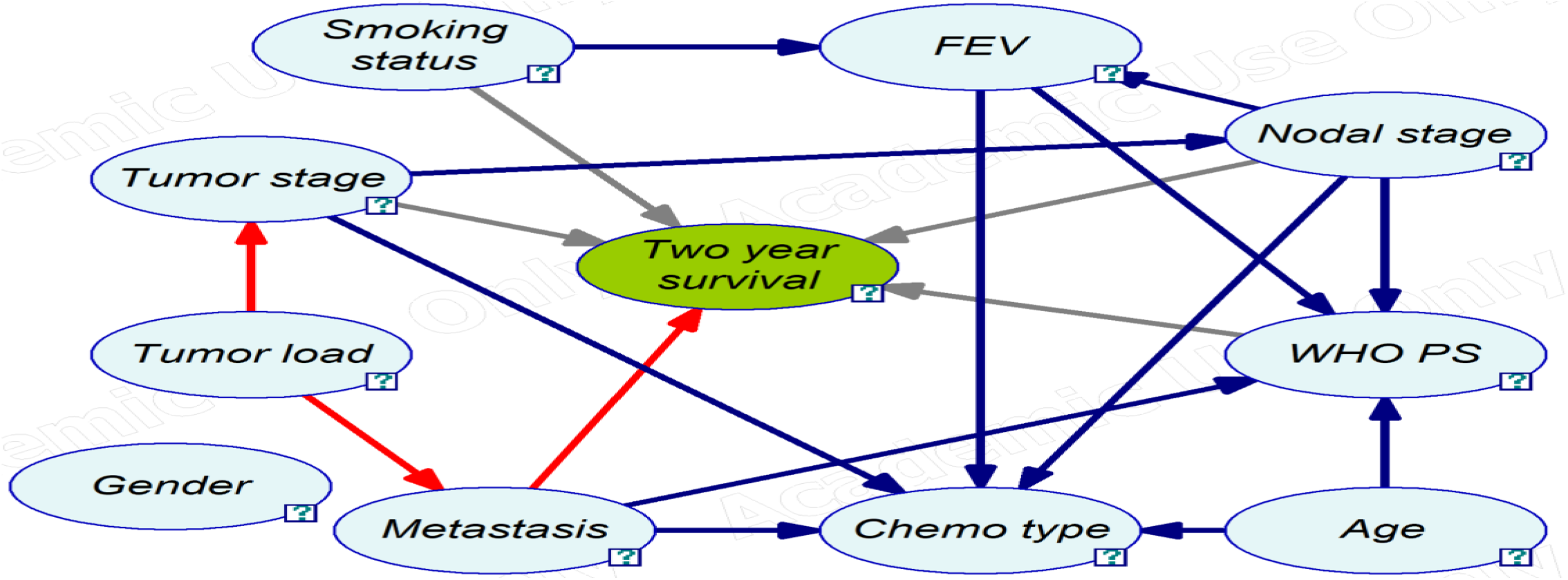
Updated Bayesian network structure to predict two-year survival with the structural.em function in the bnlearn package. The oval structure represents the variables (Node), and the arrows indicate the direction of the causal-effect relationships. Grey arrows indicate a direct parental link to the outcome of interest and red arrows indicate additional arcs. **FEV**: Forced expiratory volume, **Chemo**: Chemotherapy, **WHO PS**: World Health Organization performance status

**Fig 4.**
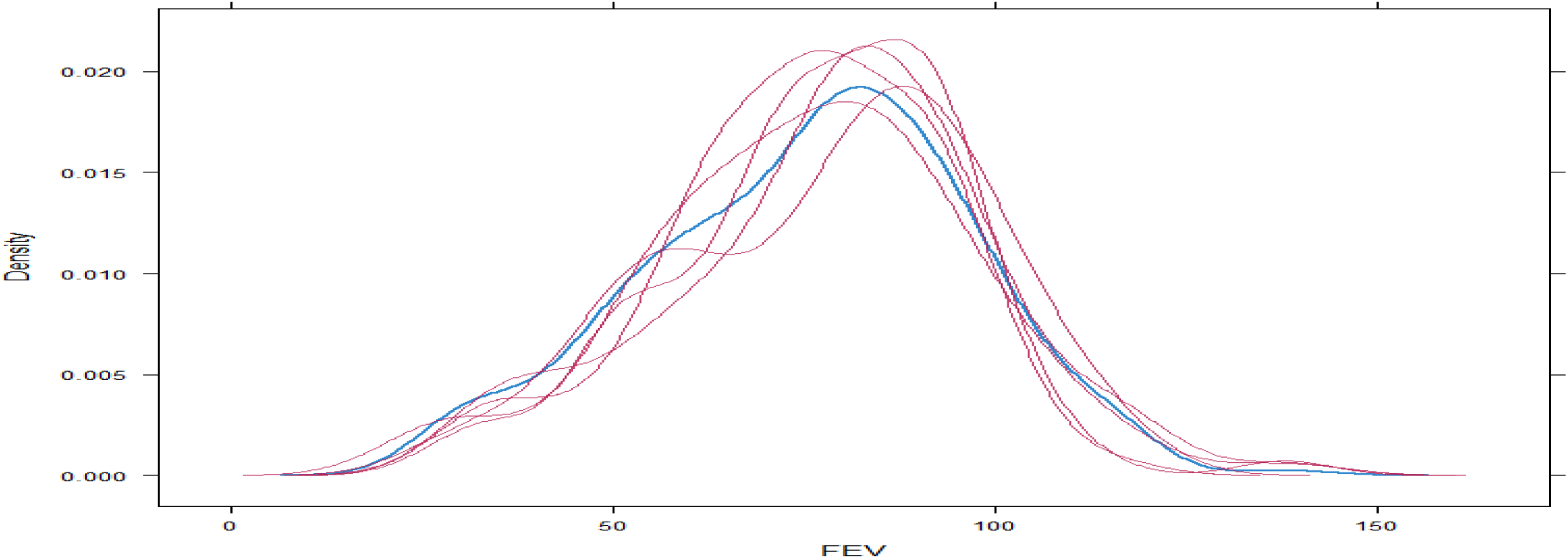
Kernel density estimates for the observed data (blue) and the imputed data (thin red lines) for forced expiratory volume (FEV).

**Fig 5.**
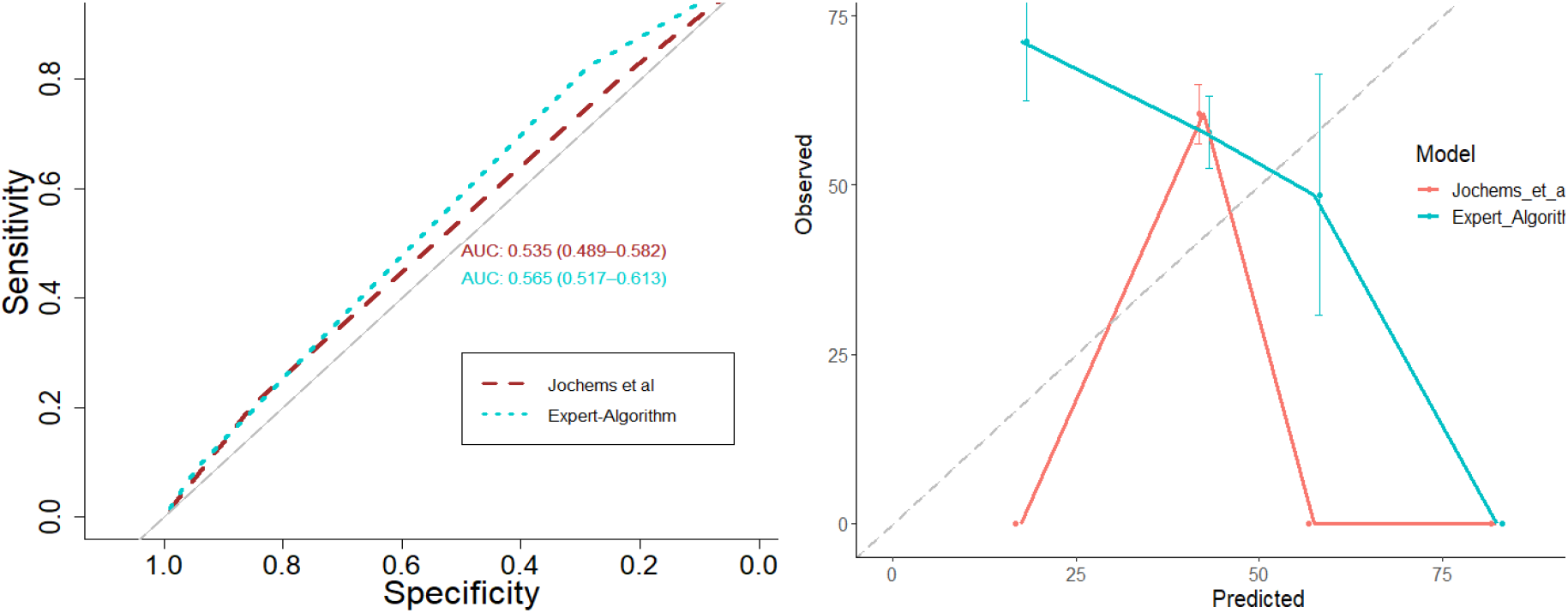
Performance of the adapted and expert-algorithm structure in terms of the area under the curve and calibration respectively.

**Table 1.** Blacklist restriction of patient characteristics.

|  | Gender | WHO PS | Age | Chemo type | Tumorload | Smoking status | Tumor stage | Nodal stage | Metastasis | FEV | Two year survival |
| --- | --- | --- | --- | --- | --- | --- | --- | --- | --- | --- | --- |
| Gender | O | X | X | X | X | X | X | X | X | L | L |
| WHO PS | X | O | T | T | T | X | T | T | T | T | L |
| Age | X | L | O | L | X | X | X | X | X | X | L |
| Chemo type | X | L | T | O | T | X | T | T | T | T | L |
| Tumorload | X | L | X | L | O | X | L | L | L | L | L |
| Smoking status | X | X | X | X | X | O | X | X | X | X | L |
| Tumor stage | X | L | X | L | T | X | O | L | X | L | L |
| Nodal stage | X | L | X | L | T | X | T | O | X | L | L |
| Metastasis | X | L | X | L | T | X | X | X | O | L | L |
| FEV | T | L | X | L | T | X | T | T | T | O | L |
| Two year survival | T | T | T | T | T | T | T | T | T | T | O |

**FEV:** Forced expiratory volume, **Chemo:** Chemotherapy
**WHO PS:** World Health Organization performance status
**T:** Top determines left, **L:** Left determines top, **X:** Both direction blocked, **O:** Diagonal

**Table 2.**
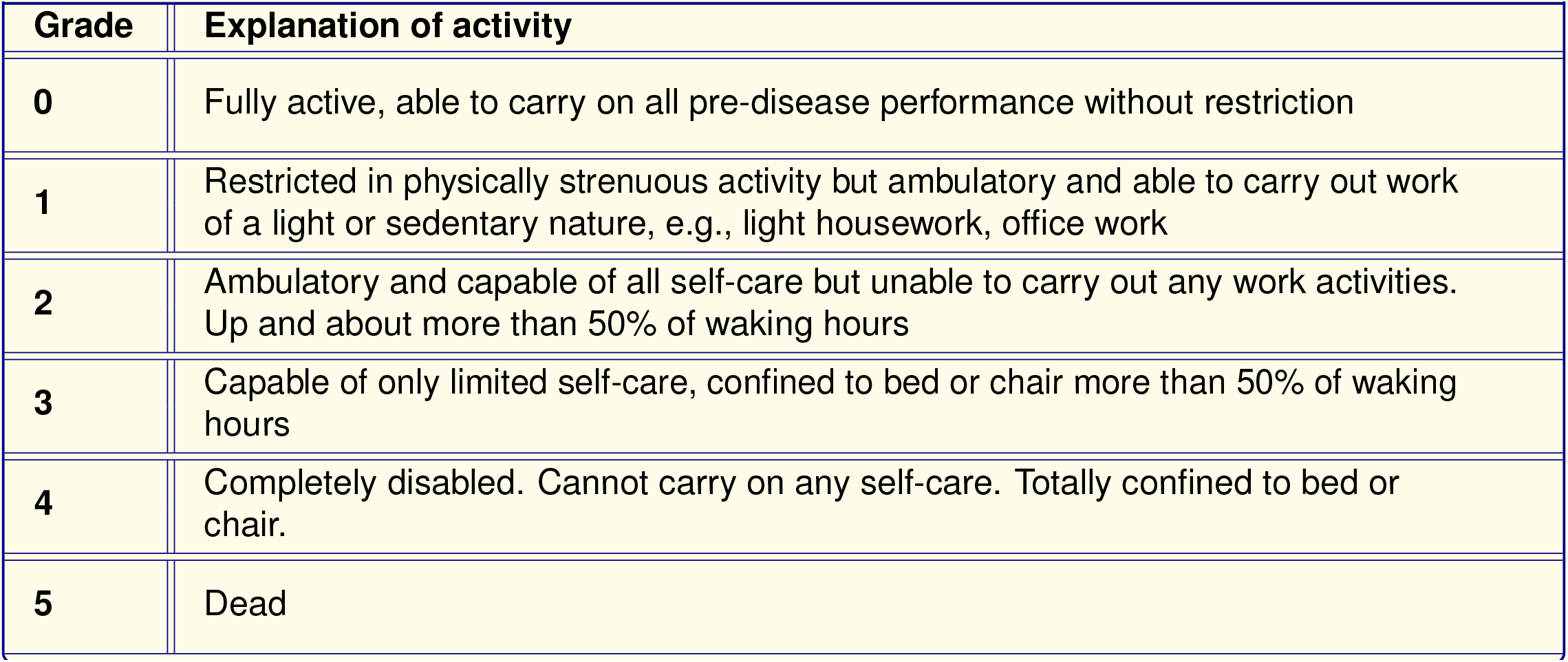
WHO Performance status

| Grade | Explanation of activity |
| --- | --- |
| **0** | Fully active, able to carry on all pre-disease performance without restriction |
| **1** | Restricted in physically strenuous activity but ambulatory and able to carry out work of a light or sedentary nature, e.g., light housework, office work |
| **2** | Ambulatory and capable of all self-care but unable to carry out any work activities. Up and about more than 50% of waking hours |
| **3** | Capable of only limited self-care, confined to bed or chair more than 50% of waking hours |
| **4** | Completely disabled. Cannot carry on any self-care. Totally confined to bed or chair. |
| **5** | Dead |

